# Inpatient treatment for severe obesity: a retrospective cohort study in Brazil

**DOI:** 10.1101/2024.04.28.24306514

**Authors:** Domingos L. S. Rios, Márcia C. A. M. Oliveira, Sérgio Q. Braga, Matheus J. Chamorro, Beatriz L. B. Cunha, Ana C. R. Reis, Ana P. Guimarães, Ana D. N. Silva, Dandara A. R. Silva, Edilene M. Q. Araújo, Magno M. W. Pimentel

## Abstract

**Introduction:** Very low-calorie diets with hospitalization have demonstrated promise as a viable therapeutic option for severe obesity and its associated comorbidities. However, large studies providing a comprehensive longitudinal observation of patients undergoing this therapy are lacking. We evaluated the effectiveness of treating severe obesity in hospitalized patients, using very low-calorie diets and clinical support to develop lifestyle changes.

**Methods:** A retrospective cohort study with a pre-post quasi-experimental design analyzed secondary data from 2016–2022 medical records of patients with severe obesity (grade II or III) treated in a Brazilian obesity specialist hospital. The patients underwent a very low-calorie diet (500–800 kCal/day) and immersive changes in lifestyle habits, monitored by a multidisciplinary team. At 3 months, 777 patients presented complete data and 402 presented complete data at 6 months. The study compared changes in bioimpedance and laboratory tests, between men and women and age groups (elderly vs. non-elderly).

**Results:** Three months of hospitalization yielded significant reductions in weight, body mass index (BMI), body fat, skeletal muscle mass, glucose, inflammatory, and lipid parameters. These reductions were more pronounced after 6 months, nearly doubling those observed at 3 months. In women, BMI and fat mass reduced by 10.4% and 15.2% at 3 months and 20.4% and 31.3% at 6 months, respectively. In men, BMI and fat mass decreased by 12.9% and 25.3 at 3 months and 23.6% and 45.3% at 6 months, respectively. Elderly individuals (aged ≥ 60 years) had smaller reductions in BMI and fat mass than non-elderly individuals (aged < 60 years) but still presented significant improvements.

**Conclusion:** This study suggests the viability of treating severe obesity by hospitalization with low-calorie diets and immersive lifestyle changes. This treatment modality significantly improves anthropometric measurements, glucose, lipids, and inflammatory markers, thereby reducing cardiovascular risk.

## Introduction

Obesity is a clinical condition characterized by increased body weight resulting from excessive fat accumulation, influenced by genetic, biological, environmental, and sociocultural factors [1].

Considered a public health problem, obesity is associated with the development of several chronic non-communicable diseases, including metabolic syndrome, dyslipidemia, insulin resistance, cardiovascular diseases, type 2 diabetes mellitus, osteoarthritis, sleep apnea, infertility, cancer, and psychological problems [2–3]. The global prevalence of obesity has increased, partly due to changes in eating patterns, with increased consumption of calorie-rich foods and unhealthy lifestyle habits, such as sedentarism.

Very low-calorie diets (VLCDs) have been proposed as a valid treatment option for grade II and III obesity, with or without comorbidities [4]. This diet involves altering and optimizing energy metabolism, stimulating the production of ketone bodies by the liver through the breakdown of fat, inducing weight loss, and improving other parameters such as insulin sensitivity and glycemic control [5–6].

Owing to the multiple interactions of causal factors, treating obesity requires an interdisciplinary and integrated therapeutic approach focused on lifestyle changes. In this scenario, hospitalization emerges as an interesting therapeutic strategy, offering a multidisciplinary intervention that successfully achieves weight loss [7].

This study aimed to evaluate the effectiveness of treating severe obesity (grades II and III) in hospitalized patients, using VLCD and clinical support to develop lifestyle changes.

## Materials and Methods

### Type of study-sample design

This retrospective cohort study was conducted using secondary data from the medical records of patients hospitalized in a controlled environment for weight loss from October 2016 to October 2022. It employed a quasi-experimental design with an interventional before-after (pre-post) design.

The research was conducted at a Brazilian hospital specialized in obesity treatment. The sample size was convenience-based, initially comprising 1,151 individuals with severe obesity hospitalized for 3 and/or 6 months. The inclusion criteria were age over 12 years, obesity grade II or III upon admission, and at least 3 months of inpatient treatment. We excluded 293 patients without data from laboratory tests and/or bioimpedance. We included 858 patients in the analysis. Of these, 321 presented complete data at 3 and 6 months, 456 presented complete data at 3 months alone, and 81 presented complete data at 6 months alone. Therefore, the analysis included 777 patients with data available at 3 months and 402 with data available at 6 months. Data was imported directly from the electronic medical records of each patient into an Excel table and then converted directly into SPSS format using SPSS software ver. 29.0.1.0 (IBM corporation, New York, USA) for statistical analysis. Data was assessed from electronic medical records from July 31, 2023 for research purposes. The authors had no access to information that could identify individual participants during and after data collection.

### Therapeutic interventions and multi-professional support

The treatment involved a multidisciplinary approach with key components including low-calorie diets (LCDs) and VLCDs, physical activity, individual cognitive behavioral therapy (CBT), participation in educational groups aimed at lifestyle changes, and multidisciplinary clinical support.

Patients received LCDs (800 to 1,200 kCal/day) or VLCDs (less than 800 kCal/day) during hospitalization. Both diets provided a higher percentage of protein (70 to 100 g/day or 0.8 to 1.5 g /kg of ideal body weight/day) and a low carbohydrate content. These diets were supplemented with vitamins, minerals, electrolytes, and essential fatty acids to ensure adequate nutrition, following Brazilian guidelines for treating obesity [8].

Patients engaged in lower-impact physical activities due to frequent arthropathies, including water aerobics exercises at least thrice a week, horizontal biking twice a week with a light load, and weight training thrice a week.

The patients underwent individual one-hour CBT sessions with a psychologist twice weekly. Patients also participated in educational group activities, using active methodologies and experiences aimed at lifestyle changes, consisting of daily one-hour meetings [9–10].

Daily clinical evaluations were conducted, with periodic consultations by endocrinologists, cardiologists, orthopedists, and psychiatrists. A team of physiotherapists, occupational therapists, and nurses assessed and monitored each patient’s condition, providing necessary interventions in their respective areas.

### Bioimpedance and anthropometry

All patients had their weight and height measured and underwent bioimpedance testing upon admission and at 3 and/or 6 months. Height was measured using a stadiometer (Tonelli Medical Devices, Brazil), and weight was measured using a bioelectrical impedance device. Body composition was assessed using a 3-frequency bioelectrical impedance device (5kHz, 50kHz, and 5,00kHz–Ottoboni-inbody570), utilizing a tetra polar system with eight points (tactile electrodes) to obtain 15 impedance measurements of each of the five body segments (right arm, left arm, trunk, right leg, and left leg). Obesity severity was assessed using body mass index (BMI): grade I, if BMI was between 30 and 34.9 kg/m^2^; grade II, between 35 and 39.9 kg/m^2^; and grade III, ≥40 kg/m^2^.

### Laboratory tests

Measurements of gamma-glutamyl transferase (GGT), blood glucose, triglycerides, total cholesterol, and high-density cholesterol (HDL) were performed using the enzymatic colorimetric method; low-density cholesterol (LDL) was calculated using the Friedewald formula. Serum zinc levels were measured using the flame atomic absorption spectrometry method. Electrochemiluminescence was used to measure ferritin and basal insulin levels. Oxaloacetic (GOT) and pyruvic (GPT) transaminases were measured using the UV-kinetic method; creatine phosphokinase (CPK) by the UV method; glycated hemoglobin HbA1c by turbidimetric inhibition immunoassay; C-reactive protein (CRP) by immunoturbidimetry. All tests were performed using an automated device and in the same laboratory.

### Statistical analysis

The bioimpedance parameters were described as median and inter-quartil interval. To compare values measured at admission with those observed after 3 and 6 months of hospitalization it was used the Wilcoxon test. The percentage change in each bioimpedance parameter was calculated by subtracting the values measured at admission from those obtained at 3 and 6 months of hospitalization, respectively. This difference was divided by the original hospitalization value and multiplied by 100. These percentage values were compared between men and women and between elderly (≥60 years) and non-elderly patients (<60 years) using a Mann-Whitney test. Laboratory measurements on admission and at 3 and 6 months of hospitalization were compared using a Wilcoxon test. Kaplan-Meier curves were used to compare the time to reach 20% weight loss and 35% fat mass reduction between male and female and between elderly and no elderly patients. To evaluate the factors associated with the success to reach median values of fat mass loss at 3 and 6 months of hospitalization, a multivariate logistic regression was performed: the dependent variables were the success of reach the median percentage of fat mass loss in each period of hospitalization (17% and 35% at 3 and 6 months hospitalization, respectively). The independent variables were included in the model, using the “forward conditional” method, the initial following dummy variables were used in the analysis: age ≥ 60 years, male sex, Diabetes Mellitus, current smoking, drinking habits, hypothyroidism, hepatic steatosis, sedentary life style and altered admission levels of glycated hemoglobin (> 5.6%), zinc (< 69.93 µg/dL), CRP (> 6 mg/L), and CPK (> 200 U/L). A significance level of 5% was adopted.

### Ethical aspects

This research was conducted following the principles of bioethics in accordance with resolution 466/2012 (CONEP/Brazil) and met aspects related to the Declaration of Helsinki. The Research Ethics Committee of the State University of Bahia (UNEB) approved the project with CAAE number 65578822.1.0000.0057. Informed consent paperwork could not be obtained due to the retrospective design of this analysis; patients were no longer available to sign informed consent. The Research Ethics Committee approved this.

## Results

Table 1 presents the clinical and epidemiological data. Women were the majority (70%) with a median age of approximately 44 years, not differing significantly between the groups by length of hospitalization. Regarding lifestyle habits, smoking (3 months: 22.1% vs. 6 months: 23.4%), drinking habits (3 months: 50.6% vs. 6 months: 49.3%) and sedentary lifestyle (3 months: 82.5% vs. 6 months: 85.6%) were prevalent and similar between the two groups.

**Table 1.** Clinical and epidemiological characteristics.

| Variable | Three months<br>tratment | Six months<br>treatment | p* |
| --- | --- | --- | --- |
|  | (n=777) | (n=402) |  |
| **Female sex** | 70.3% (546) | 70.6% (284) | 0.947 |
| **Age (years)** | 44 (33-59) | 46 (34-61,2) | 0.102** |
| **Current smoking** | 22.1% (172) | 23.4% (94) | 0.680 |
| **Drinking habits** | 50.6% (393) | 49.3% (198) | 0.711 |
| **Sedentary lifestyle** | 82.5% (641) | 85.6% (344) | 0.205 |
| **Diabetes Mellitus** | 34.6% (269) | 32.6% (131) | 0.526 |
| **Hypertension** | 53% (412) | 52% (209) | 0.783 |
| **Hypercholesteroleamia** | 44.3% (344) | 42.8% (172) | 0.670 |
| **Hypertriglyceridemia** | 30.9% (240) | 31.1% (125) | 0.995 |
| **Hypothyreiodism** | 15.3% (119) | 17.4% (70) | 0.397 |
| **Coronary Artery Disease** | 16% (124) | 17.4% (70) | 0.579 |
| **Sleep apnea** | 75.3% (585) | 78.1% (314) | 0.314 |
| **Hepatic steatosis** | 75.8% (589) | 74.4% (299) | 0.640 |
| **Depression \*\*\*** | 30.8% (239) | 33.1% (133) | 0.454 |
| **Obesity grade II** | 37.1% (288) | 24.1% (97) | <0.001 |
| **Obesity grade III** | 62.9% (489) | 75.9% (305) | <0.001 |

\* Pearson chi-square with Yates' correction;
\*\* MannWhitney test, data is presented as median and interquartile interval values;
\*\*\* Self-referred.

The prevalence of diabetes mellitus (3 months: 32.6% vs. 6 months: 34.6%), hypertension (3 months: 52% vs. 6 months: 53%), hypercholesterolemia (3 months: 42.8% vs. 6 months: 44.3%) and hypertriglyceridemia (3 months: 31.1% vs. 6 months: 30.9%) was high and did not differ significantly between groups by length of hospitalization. Hepatic steatosis (3 months: 74.4% and 6 months: 75.8%), sleep apnea (3 months: 78.1% and 6 months: 75.3%) and depression (3 months: 33.1% and 6 months: 30.8%) were prevalent in both periods of hospitalization. Patients with 6 months of hospitalization had a higher percentage of most severe obesity (grade III: 75.9%) than patients who remained hospitalized for 3 months (62.9%) (Table 1).

After 3 months of hospitalization, body weight reduced from 112.7kg to 100.2kg, BMI from 40.9 kg/m^2^ to 36.4 kg/m^2^, fat mass from 55.7kg to 45.8kg, fat percentage from 51.3% to 47.8%, skeletal muscle mass from 31.3kg to 28.9kg, basal metabolic rate from 1,568 kCal to 1,498 kCal, and waist-to-hip ratio (WHR) from 1.07 to 1.00 (Table 2).

**Table 2.** Changes in bioimpedance based on length of hospitalization.

| Parameters | Admission<br>n = 777 | Three months<br>n = 777 | p* | Admission<br>n = 402 | Six months<br>n = 402 | p* |
| --- | --- | --- | --- | --- | --- | --- |
| Weight (kg) | 112.7 (100.9-126.3) | 100.2 (89.7-111.7) | <0.001 | 116.4 (103.7-130.2) | 91.6 (83.8-101.9) | <0.001 |
| BMI (kg/m<sup>2</sup>) | 40.9 (39.1-44.5) | 36.4 (34.7-39.4) | <0.001 | 42.6 (40.0-46.1) | 33.4 (31.5-36.4) | <0.001 |
| Fat mass (kg) | 55.7 (50.4-63.9) | 45.8 (40.6-52.3) | <0.001 | 58.9 (52.9-67.2) | 38.0 (32.9-44.9) | <0.001 |
| Body fat percentage(%) | 51.3 (48.6-53.3) | 47.8 (43.0-51.3) | <0.001 | 52.0 (50.0-53.7) | 43.2(37.6-48.0) | <0.001 |
| Skeletal muscle mass (kg) | 31.3 (27.3-36.9) | 28.9 (24.8-34.4) | <0.001 | 30.6 (27.1-36.3) | 29.0 (25.4-34.6) | <0.001 |
| Basal metabolic rate (kCal) | 1568.0 (1436.8-1775.5) | 1498.5 (1354.5-1698.8) | <0.001 | 1549.0 (1423.5-1760.0) | 1497.0 (1368.0-1697.5) | <0.001 |
| Waist to hip ratio | 1.07 (1.00-1.14) | 1.00 (0.95-1.05) | <0.001 | 1.08 (1.02-1.13) | 1.03 (0.98-1.08) | <0.001 |

\*Wilcoxon test, data is presented as median and interquartile interval values.

Patients hospitalized for 6 months demonstrated an even greater reduction in all analyzed bioimpedance parameters. Body weight reduced from 116.4kg to 91.6kg, BMI from 42.6kg/m^2^ to 33.4kg/m^2^; fat mass from 58.9kg to 38kg, fat percentage from 52% to 43.2%, skeletal muscle mass from 30.6kg to 29kg, basal metabolic rate from 1,549 kCal to 1,497 kCal, and WHR from 1.08 to 1.03 (Table 2).

After 3 months of hospitalization, men exhibited a higher percentage of reduction than women in the following bioimpedance parameters: weight (men: 12.9% vs. women: 10.4%), BMI (men: 12.9 % vs. women: 10.4%), fat mass (men: 25.3% vs. women: 15.2%), body fat percentage (men: 13.6% vs. women: 5.2%), and WHR (men: 6.5% vs. women: 3.3%). However, for basal metabolic rate (men: 2.5% vs. women: 3.8%) and skeletal muscle mass (men: 3.8% vs. women: 5.9%), men exhibited a lower reduction than women (Table 3).

**Table 3.** Changes in bioimpedance by sex.

| Parameters | Three months |  |  | Six months |  |  |
| --- | --- | --- | --- | --- | --- | --- |
|  | Female<br>n = 546 | Male<br>n = 231 | P* | Female<br>n = 284 | Male<br>n = 118 | P* |
| Weight lost percentual (%) | 10.4 (8.9-12.0) | 12.9 (11.1-14.7) | <0.001 | 20.4 (17.4-23.4) | 23.6 (20.9-27.4) | <0.001 |
| BMI lost percentual (%) | 10.4 (8.9-12.0) | 12.9 (11.1-14.7) | <0.001 | 20.4 (17.4-23.4) | 23.6 (20.9-27.4) | <0.001 |
| Fat mass lost percentual (%) | 15.2 (12.3-18.5) | 25.3 (19.6-30.0) | <0.001 | 31.3 (26.0-37.8) | 45.3 (36.0-51.6) | <0.001 |
| Reduction in body fat percentual (%) | 5.2 (3.0-8.6) | 13.6 (8.7-18.3) | <0.001 | 14.3 (8.4-20.5) | 27.3 (17.7-35.3) | <0.001 |
| Skeletal muscle mass lost percentual (%) | 5.9 (7.3-8.3) | 3.8 (1.4 - 6.4) | <0.001 | 8.7 (5.4-12.8) | 6.6 (2.3-10.2) | <0.001 |
| Basal metabolic rate reduction percentual (%) | 3.8 (1.9-5.5) | 2.5 (0.7 - 4.3) | <0.001 | 5.5 (2.9-8.5) | 4.3 (1.4-7.0) | 0.002 |
| Waist to hip ratio reduction percentual (%) | 3.3 (0-6.2) | 6.5 (3.2 - 9.6) | <0.001 | 6.2 (1.2-9.5) | 10.0 (3.8-16.6) | <0.001 |

\* MannWhitney test, data is presented as median and interquartile interval values.

Similarly, but to a greater extent, at 6 months of hospitalization, men exhibited higher percentages of loss in weight (men: 23.6% vs. women: 20.4%), BMI (men: 23.6% vs. women: 20.4%), fat mass (men: 45.3% vs. women: 31.3%), body fat percentage (men: 27.3% vs. women: 14.3%), and WHR (men: 10% vs. women: 6.2%). Conversely, women had higher rates of loss of skeletal muscle mass (men: 6.6% vs. women 8.7%) and basal metabolic rate (men: 4.3% vs. women: 5. 5%) (Table 3).

The Survival Kaplan-Meier curves compares male and female for the time to reach reductions of 20% weight (Figure 1) and 35% fat mass (Figure 2) in the period of treatment. The females reach 20% weight reduction by the fifth month while males reach this by the fourth month of treatment (p<0.001). The females reach 35% fat mass reductions by the sixth month while males reach this in the fourth month of treatment (p<0.001).

**Fig 1.**
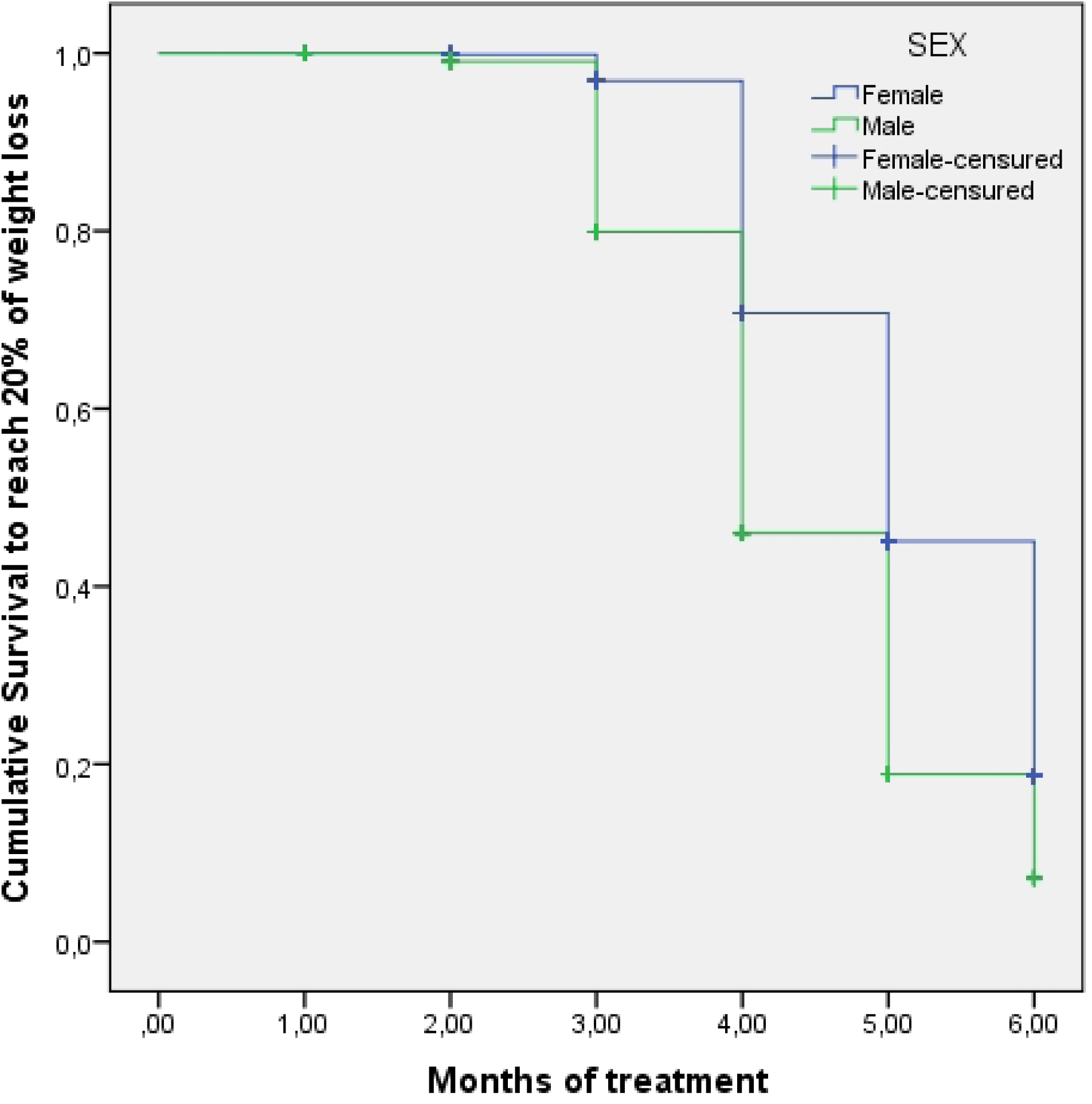
Weight loss survival curve by sex during inpatient treatment. Medians to reach 20% of weight loss: Female 5 months (95%CI: 4.8-5.2 months); Male 4 months (95%CI: 3.8-4.2 months); Log rank (Mantel-Cox test): 73.03; p<0.001.

**Fig 2.**
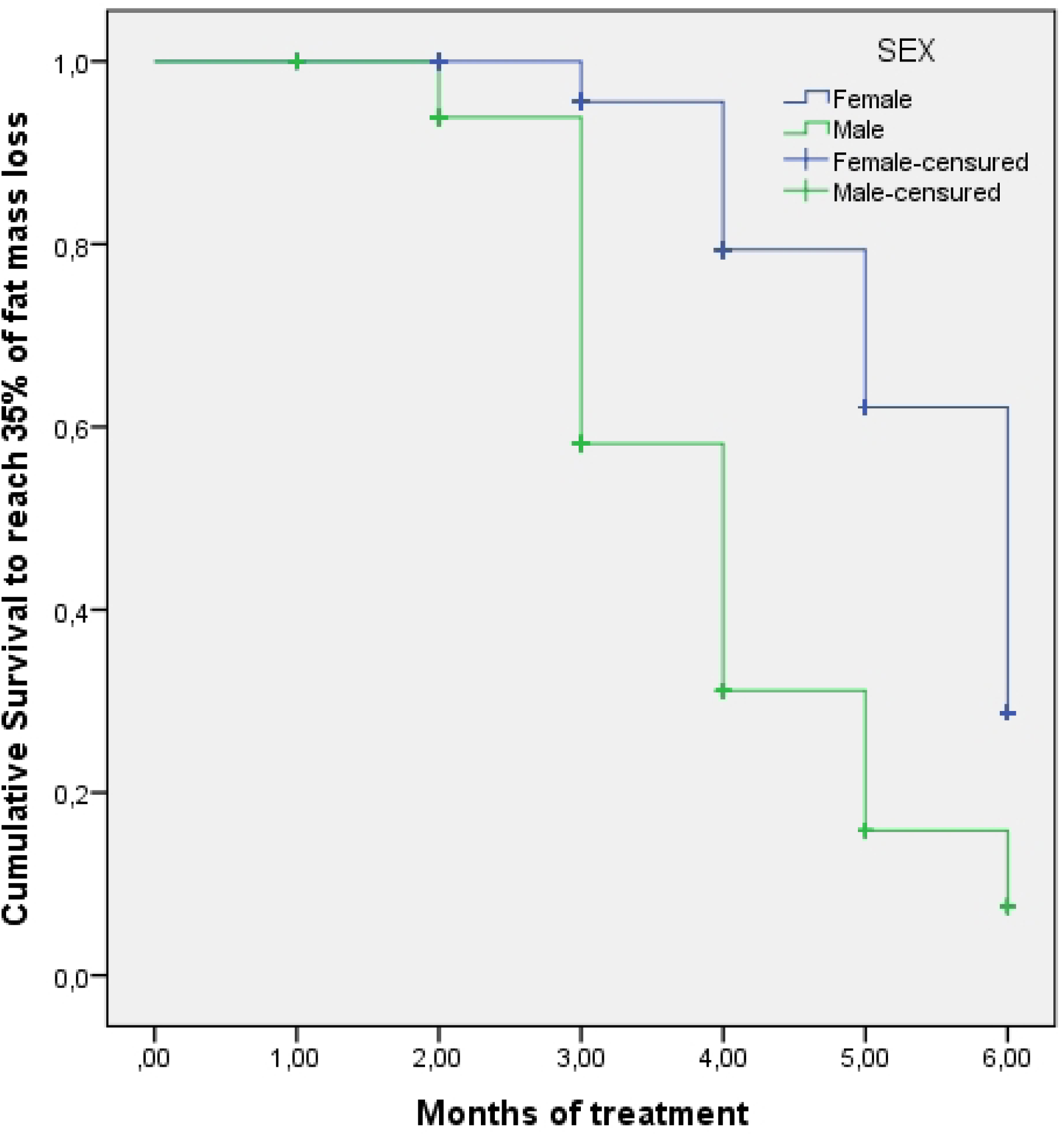
Fat mass loss survival curve by sex during inpatient treatment. Medians to reach 35% of fat mass loss: Female 6 months (95%CI: 5.8-6.2 months); Male 4 months (95%CI: 3.8-4.2 months); Log rank (Mantel-Cox test): 236.885; p<0.001.

After 3 months of hospitalization, the elderly (≥ 60 years) demonstrated a lower percentage of loss in weight (elderly: 10.2% vs. non-elderly: 11.4%), BMI (elderly: 10.3% vs. non-elderly: 11, 4%), fat mass (elderly: 14.5% vs. non-elderly: 17.6%), body fat percentage (elderly: 4.9% vs. non-elderly: 7.3%), and WHR (elderly: 3.6% vs. non-elderly: 4.4%) than the non-elderly. The percentage differences in skeletal muscle mass loss (elderly: 5.9% vs. non-elderly: 5.2%) but not in the basal metabolic rate (elderly: 3.8% vs. non-elderly: 3.3%) was higher in elderly as compared to no elderly patients (Table 4).

**Table 4.** Changes in bioimpedance by age.

| Parameters | Three months |  | p* | Six months |  | p* |
| --- | --- | --- | --- | --- | --- | --- |
|  | Non-elderly<br>n = 598 | Elderly<br>n = 179 |  | Non-elderly<br>n = 290 | Elderly<br>n = 112 |  |
| Weight lost percentual (%) | 11.4 (9.6-13.0) | 10.2 (8.4-12.1) | <0.001 | 21.8 (18.8-25.4) | 19.8 (17.0-22.7) | <0.001 |
| BMI lost percentual (%) | 11.4 (9.6-13.0) | 10.2 (8.4-12.1) | <0.001 | 21.8 (18.8-25.4) | 19.8 (17.0-22.7) | <0.001 |
| Fat mass lost percentual (%) | 17.6 (13.8-23.5) | 14.5 (11.4-19.5) | <0.001 | 37.0 (29.4-45.4) | 30.5 (24.9-35.9) | <0.001 |
| Reduction in body fat percentual (%) | 7.3 (4.1-12.4) | 4.9 (2.8-9.7) | <0.001 | 18.9 (11.9-27.1) | 12.9 (8.0-17.2) | <0.001 |
| Skeletal muscle mass lost percentual (%) | 5.2 (2.3-7.7) | 5.9 (3.7-8.1) | 0.026 | 7.2 (3.9-11.8) | 10.2 (6.6-12.8) | <0.001 |
| Basal metabolic rate reduction percentual (%) | 3.3 (1.3-5.2) | 3.8 (2.0 - 5.3) | 0.124 | 4.6 (2.3-8.0) | 6.3 (3.9-8.2) | <0.001 |
| Waist to hip ratio reduction percentual (%) | 4.4 (1.1-7.3) | 3.6 (-1.0 - 6.7) | 0.006 | 7.6 (3.0-12.3) | 4.4 (-2.0 - 8.7) | <0.001 |

\* Mann-Whitney test, data is presented as median and interquartile interval values.

Similarly, after 6 months of hospitalization, the elderly had a lower percentage of loss in weight (elderly: 19.8% vs. non-elderly: 21.8%), BMI (elderly: 19.8% vs. non-elderly: 21.8%), fat mass (elderly: 30.5% vs. non-elderly: 37.0%), body fat percentage (elderly: 12.9% vs. non-elderly: 18.9%) and WHR (elderly: 4.4% vs. non-elderly: 7.6%). Contrary to the observation after 3 months, elderly individuals had a higher percentage of loss of skeletal muscle mass (elderly: 10.2% vs. non-elderly: 7.2%) and basal metabolic rate (elderly: 6.3 % vs. non-elderly: 4.6%) at 6 months of hospitalization (Table 4).

Three months of hospitalization yielded a significant reduction in the levels of fasting glucose (admission: 93.0 mg/dL vs. 3 months: 88.0 mg/dL), insulin (admission: 22.2 µIU/mL vs. 3 months: 16.8 µIU/mL) and glycated hemoglobin (admission: 5.6% vs. 3 months: 5.3%); triglycerides (admission: 119.0 mg/dL vs. 3 months: 94.0 mg/dL), HDL (admission: 45 mg/dL vs. 3 months: 43 mg/dL), LDL (admission: 114 mg/dL vs. 3 months: 90 mg/dL), and total cholesterol (admission: 185 mg/dL vs. 3 months: 153 mg/dL).

Regarding liver injury and inflammatory markers, CRP (admission: 8.2 mg/L vs. 3 months: 5.2 mg/dL), ferritin (admission: 165 ng/mL vs. 3 months: 137 ng/mL), and GGT (admission: 35 mg/dL vs. 3 months: 25 mg/dL) levels significantly reduced after 3 months of treatment. However, GPT increased slightly (admission: 24.5 mg/dL vs. 3 months: 27.2 mg/dL) while GOT remained unchanged during this period of hospitalization (Table 5).

**Table 5.**
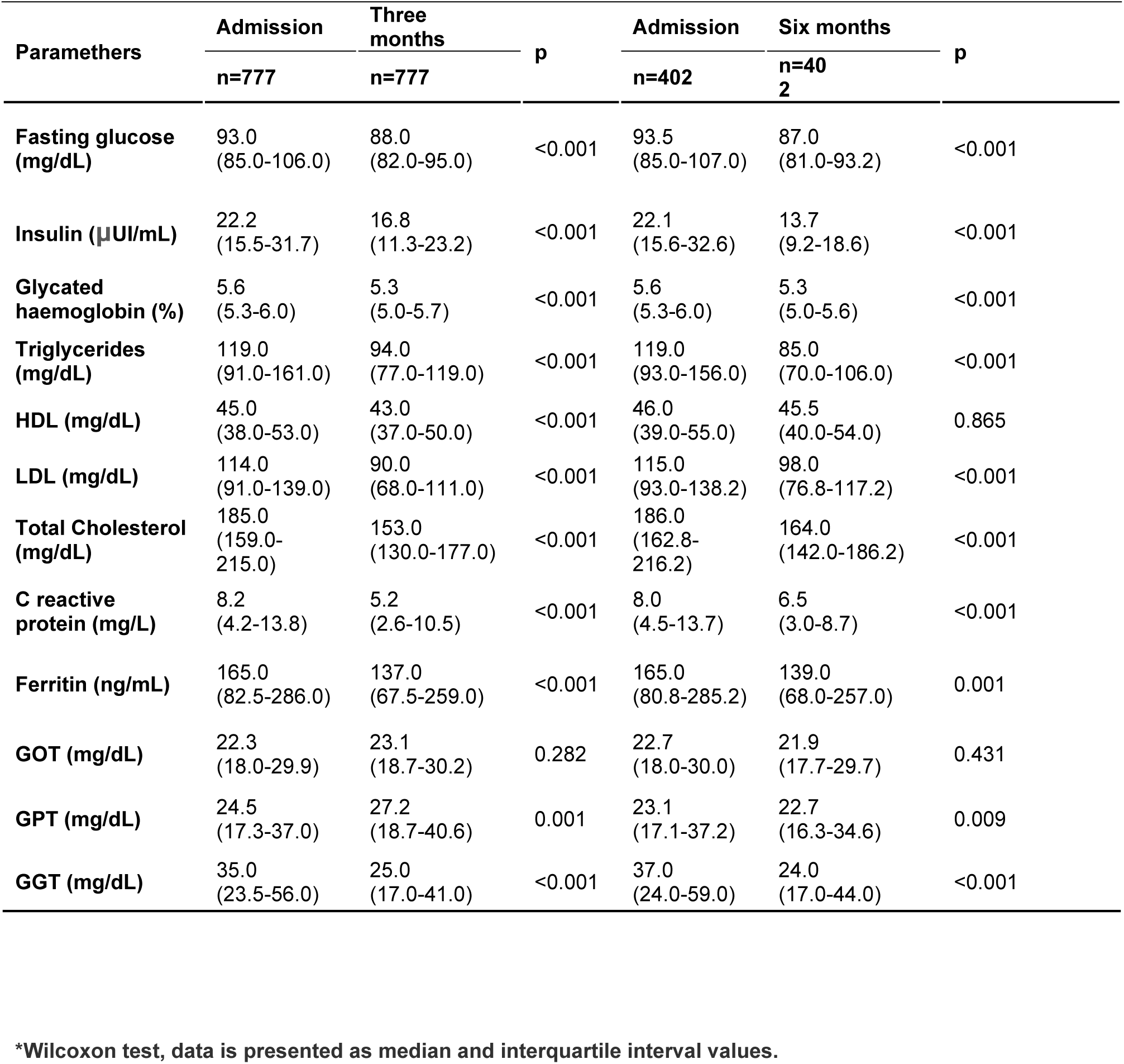
Changes* in glycide, lipid, and hepatic parameters.

| Parameters | Admission | Three months | p | Admission | Six months | p |
| --- | --- | --- | --- | --- | --- | --- |
|  | n=777 | n=777 |  | n=402 | n=402 |  |
| **Fasting glucose (mg/dL)** | 93.0<br>(85.0-106.0) | 88.0<br>(82.0-95.0) | <0.001 | 93.5<br>(85.0-107.0) | 87.0<br>(81.0-93.2) | <0.001 |
| **Insulin (μUI/mL)** | 22.2<br>(15.5-31.7) | 16.8<br>(11.3-23.2) | <0.001 | 22.1<br>(15.6-32.6) | 13.7<br>(9.2-18.6) | <0.001 |
| **Glycated haemoglobin (%)** | 5.6<br>(5.3-6.0) | 5.3<br>(5.0-5.7) | <0.001 | 5.6<br>(5.3-6.0) | 5.3<br>(5.0-5.6) | <0.001 |
| **Triglycerides (mg/dL)** | 119.0<br>(91.0-161.0) | 94.0<br>(77.0-119.0) | <0.001 | 119.0<br>(93.0-156.0) | 85.0<br>(70.0-106.0) | <0.001 |
| **HDL (mg/dL)** | 45.0<br>(38.0-53.0) | 43.0<br>(37.0-50.0) | <0.001 | 46.0<br>(39.0-55.0) | 45.5<br>(40.0-54.0) | 0.865 |
| **LDL (mg/dL)** | 114.0<br>(91.0-139.0) | 90.0<br>(68.0-111.0) | <0.001 | 115.0<br>(93.0-138.2) | 98.0<br>(76.8-117.2) | <0.001 |
| **Total Cholesterol (mg/dL)** | 185.0<br>(159.0-215.0) | 153.0<br>(130.0-177.0) | <0.001 | 186.0<br>(162.8-216.2) | 164.0<br>(142.0-186.2) | <0.001 |
| **C reactive protein (mg/L)** | 8.2<br>(4.2-13.8) | 5.2<br>(2.6-10.5) | <0.001 | 8.0<br>(4.5-13.7) | 6.5<br>(3.0-8.7) | <0.001 |
| **Ferritin (ng/mL)** | 165.0<br>(82.5-286.0) | 137.0<br>(67.5-259.0) | <0.001 | 165.0<br>(80.8-285.2) | 139.0<br>(68.0-257.0) | 0.001 |
| **GOT (mg/dL)** | 22.3<br>(18.0-29.9) | 23.1<br>(18.7-30.2) | 0.282 | 22.7<br>(18.0-30.0) | 21.9<br>(17.7-29.7) | 0.431 |
| **GPT (mg/dL)** | 24.5<br>(17.3-37.0) | 27.2<br>(18.7-40.6) | 0.001 | 23.1<br>(17.1-37.2) | 22.7<br>(16.3-34.6) | 0.009 |
| **GGT (mg/dL)** | 35.0<br>(23.5-56.0) | 25.0<br>(17.0-41.0) | <0.001 | 37.0<br>(24.0-59.0) | 24.0<br>(17.0-44.0) | <0.001 |

\*Wilcoxon test, data is presented as median and interquartile interval values.

Similarly, but to a greater extent, after 6 months of hospitalization, almost all parameters evaluated reduced significantly, including fasting glucose (admission: 93.5 mg/dL vs. 6 months: 87.0 mg/dL); insulin (admission: 22.1 µIU/mL vs. 6 months: 13.7 µIU/mL); glycated hemoglobin (admission: 5.6% vs. 6 months: 5.3%); triglycerides (admission: 119 mg/dL vs. 6 months: 85 mg/dL); LDL (admission: 115 mg/dL vs. 6 months: 98 mg/dL), and total cholesterol (admission: 186 mg/dL vs. 6 months: 164 mg/dL). However, HDL levels remained unchanged (admission: 46 mg/dL vs. 6 months: 45.5 mg/dL). Regarding liver injury and inflammatory parameters, a significant reduction was observed in the values of GGT (admission: 37 mg/dL vs. 6 months: 24 mg/dL), GPT (admission: 23.1 mg/dL vs. 6 months: 22.7 mg/dL), CRP (admission: 8 mg/L vs. 6 months: 6.5 mg/L), and ferritin (admission: 165 ng/mL vs. 6 months: 139 ng/mL). Contrary to the observation at 3 months, GOT levels increased within the normal range (admission: 22.7 mg/dL vs. 6 months: 21.9 mg/dL) (Table 5).

The Survival Kaplan-Meier curves compares non-elderly and elderly patients for the time to reach reductions of 20% in weight (Figure 3) and 35% in fat mass (Figure 4) during treatment. Elderly patients reach 20% weight (p<0.001) and 35% fat mass (p<0.001) reductions by the sixth month of treatment as compared to 5 months for non-elderly patients.

**Fig 3.**
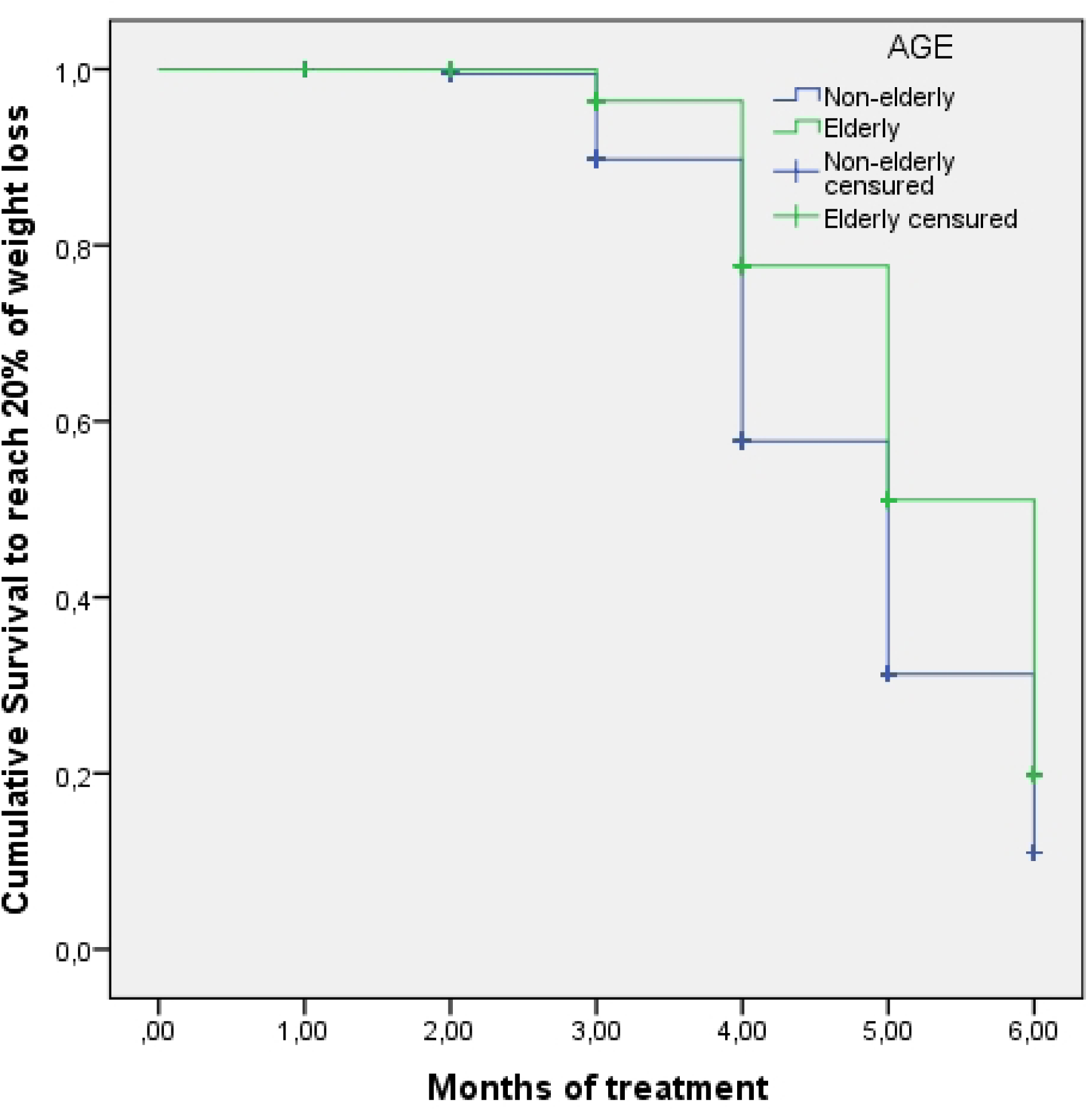
Weight loss survival curve by age during inpatient treatment. Medians to reach 20% of weight loss: Non-elderly patients 5 months (95%CI: 4.8-5.2 months); Elderly patients 6 months (95%CI: 5.8-6.2 months); Log rank (Mantel-Cox test): 29.47; p<0.001.

**Fig 4.**
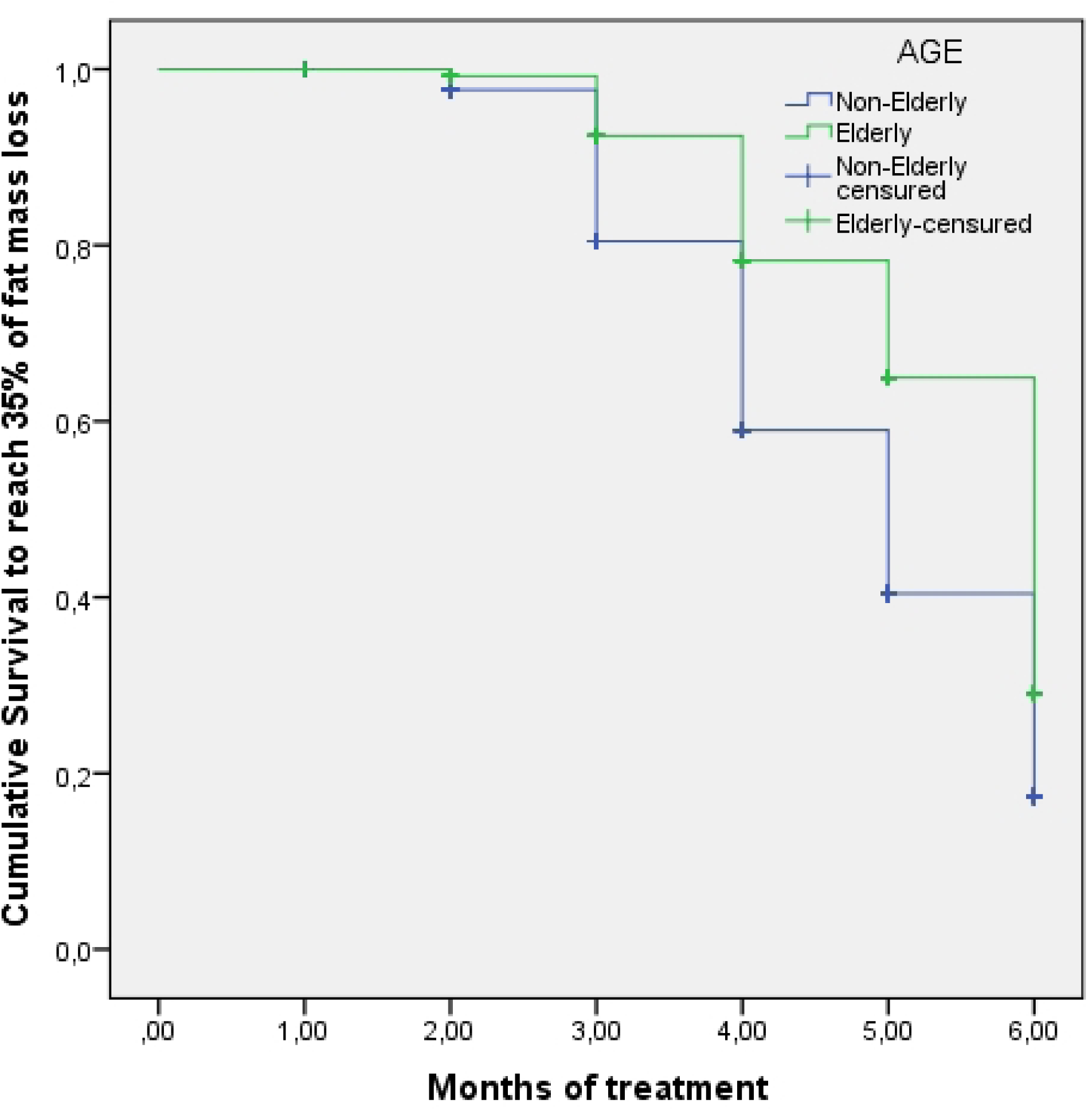
Fat mass loss survival curve by age during inpatient treatment. Medians to reach 35% of fat mass loss: Non-elderly patients 5 months (95%CI: 4.8-5.2 months); Elderly patients 6 months (95%CI: 5.8-6.2 months); Log rank (Mantel-Cox test): 29.465; p<0.001.

Table 6 presents the multivariate logistic regression with the factors, measured at admission, associated with the success to reach the median percentage of fat mass loss during hospitalization. After three months, male sex, drinking habits, CPK above normal range (>200 U/L) were associated with a higher odds ratio to reach the median percentage of body fat mass loss (17%). An inverse association was observed with elderly (age ≥ 60 years), C-reactive protein level above the normal range (> 6mg/dl), and zinc level below the normal range (< 69,93 µg/dL) [Table 6].

**Table 6.**
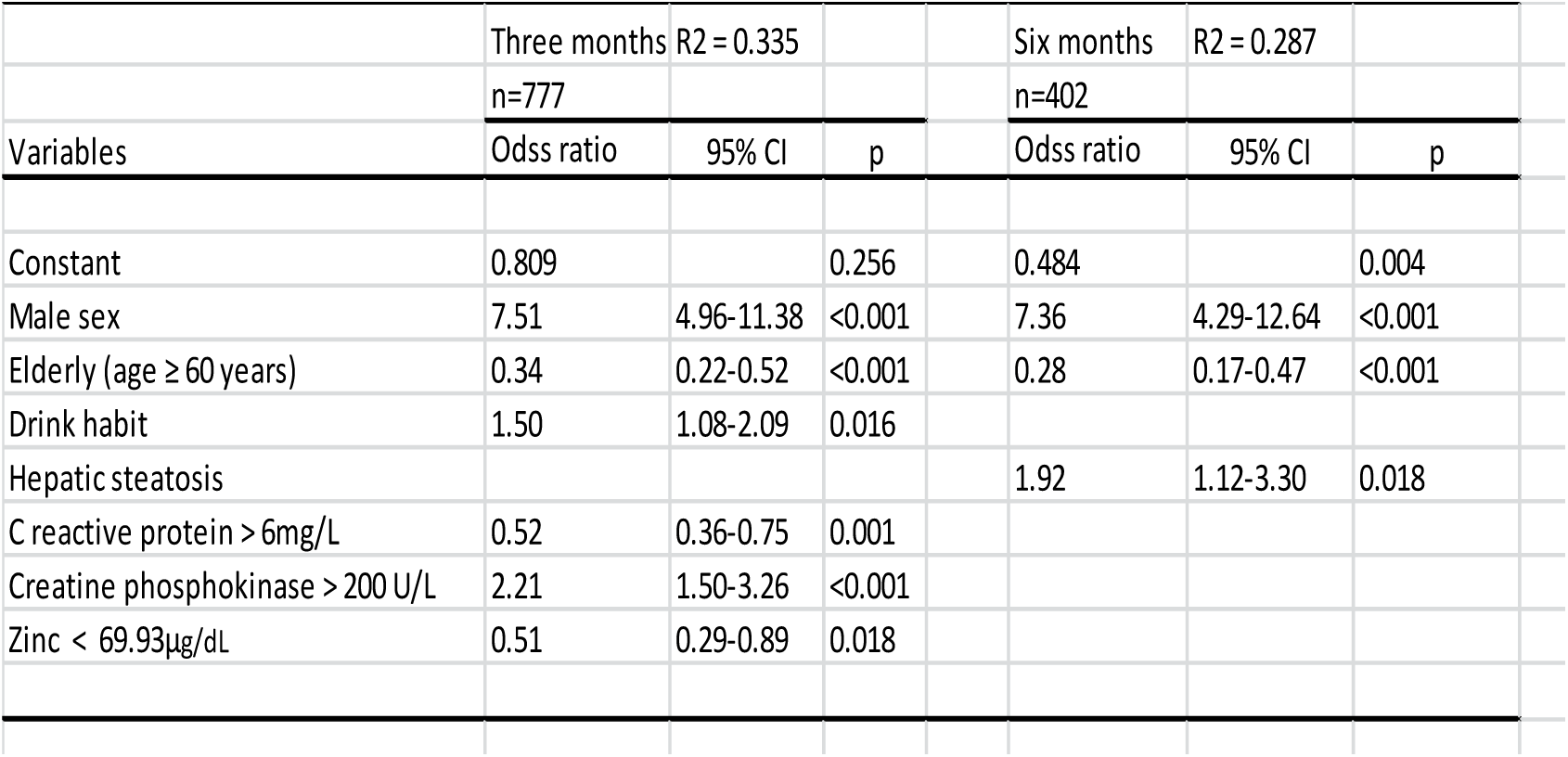
Factor associated with body fat mass loss* in both periods of inpatient treatment.

|  | Three months R2 = 0.335 |  |  |  | Six months R2 = 0.287 |  |  |
| --- | --- | --- | --- | --- | --- | --- | --- |
|  | n=777 |  |  |  | n=402 |  |  |
| Variables | Odss ratio | 95% CI | p |  | Odss ratio | 95% CI | p |
| Constant | 0.809 |  | 0.256 |  | 0.484 |  | 0.004 |
| Male sex | 7.51 | 4.96-11.38 | <0.001 |  | 7.36 | 4.29-12.64 | <0.001 |
| Elderly (age ≥ 60 years) | 0.34 | 0.22-0.52 | <0.001 | | 0.28 | 0.17-0.47 | <0.001 |
| Drink habit | 1.50 | 1.08-2.09 | 0.016 |  |  |  |  |
| Hepatic steatosis |  |  |  |  | 1.92 | 1.12-3.30 | 0.018 |
| C reactive protein > 6mg/L | 0.52 | 0.36-0.75 | 0.001 |  |  |  |  |
| Creatine phosphokinase > 200 U/L | 2.21 | 1.50-3.26 | <0.001 |  |  |  |  |
| Zinc < 69.93μg/dL | 0.51 | 0.29-0.89 | 0.018 | | | | |

At six months of hospitalization, some of these predictors lost significance, with male sex, hepatic steatosis, and elderly remaining significant. Similar to the results at 3 months, elderly people presented a lower odds ratio to reach the median percentage of body fat mass loss at 6 months (34%), while males presented higher odds ratio to reach such loss. Alcohol consumption and glycated hemoglobin levels, testosterone, zinc, and magnesium were no longer significant predictors of body fat mass loss at 6 months (Table 6).

## Discussion

Most hospitalized patients were women, presented with drinking habits, and had a sedentary lifestyle. The most prevalent comorbidities were hypertension, diabetes, hypercholesterolemia, hypertriglyceridemia, hypothyroidism, coronary artery disease, sleep apnea, hepatic steatosis, and self-reported depression. These data did not differ between the 3 and 6-month hospitalization groups. The group that underwent 6 months of treatment had a higher percentage of patients with grade III obesity, which partly justifies the longer hospital stay in this group. In a study of patients with obesity grade III who underwent outpatient dietary treatment in Rio de Janeiro, Brazil, most participants were women (76.3%) and adults (average age of 45 years), similar to this study. However, there was a higher prevalence of hypertension (80%) and diabetes (46%) and a lower incidence of drinking habits (37%), sleep apnea (31%), and dyslipidemia (16%) [11], compared to the patient profile of this study. This study is based on a Brazilian, mixed population of predominantly African (50.5%) and European (42.4%) ancestry and fewer Native Americans (5.8%) [12].

Both groups revealed significant changes in body composition parameters throughout each period, with a more significant improvement in these variables after 6 months of hospitalization.

After 3 months, we observed a significant decrease in weight, BMI, fat mass, body fat percentage, skeletal muscle mass, basal metabolic rate, and WHR. These results indicate a positive response to short-term treatment, suggesting the effectiveness of hospitalization in promoting weight loss and improving body composition. This finding aligns with a study [13] that investigated the effects of short-term multidisciplinary interventions in patients with obesity and demonstrated significant reductions in fat mass, percentage of body fat, and BMI. Skeletal muscle mass and basal metabolic rate also reduced significantly but at a magnitude approximately six times smaller than the fat mass loss (1.6 kg vs. 10.9 kg, respectively), even at six months of treatment.

Patients hospitalized for 6 months demonstrated more significant changes in body composition measured by bioimpedance. The magnitude of these changes, especially in the loss of weight (men: 23.6% and women: 20.4%) and fat mass (men: 45.3% and women: 31.3%), suggests that 6 months of hospitalization doubles the reduction in these parameters, with an eight times smaller loss of skeletal muscle mass.

In both periods, men exhibited higher percentages of loss in weight, fat mass, percentage of body fat, and WHR, in addition to lesser loss of skeletal muscle mass compared to women. These results corroborate previous findings demonstrating that in short periods of intervention, men experience a more pronounced response in weight reduction with greater preservation of skeletal muscle mass [14–15].

In addition to the differences in bioimpedance parameters previously discussed, the unequal reduction in WHR between men and women was notable in both periods of hospitalization. Men demonstrated a significantly greater reduction (almost double) in WHR. This observation is consistent with previous findings that men have a more pronounced loss of visceral fat and, consequently, a greater reduction in WHR [16–17].

Reduced WHR is associated with cardiovascular benefits and reduced health-related adverse events. Epidemiological studies highlight that a lower WHR reduces the risk of cardiovascular diseases, hypertension, and type 2 diabetes [18]. This association lies in the direct relationship between WHR and visceral fat and, consequently, with insulin resistance and inflammation caused by excess of this fat, factors that play crucial roles in increasing the risk of cardiovascular diseases [19–20]. Despite the different reduction levels in WHR, both sexes improved these measures, suggesting that both men and women experience the benefits in cardiovascular risk resulting from these reductions.

Regarding elderly and non-elderly patients, a reduction in weight, BMI, fat mass, percentage of body fat mass, and WHR was observed in both periods. However, non-elderly patients presented better results than elderly patients. A previous study [21] also evaluated the repercussions of treating severe obesity in an inpatient setting in elderly and non-elderly patients. Their results are consistent with ours because although both groups obtained significant results, non-elderly patients exhibited a greater BMI, weight, and waist circumference reduction than the elderly patients.

The aging process naturally modifies body composition, which can lead to an increase of 2 to 5% in total fat mass each decade from the age of 40, in addition to a reduction in muscle mass [22]. In this study, the differences in the loss of skeletal muscle mass and reduction in basal metabolic rate were not significant between age groups during the 3-month hospitalization. However, during the 6-month hospitalization, elderly patients exhibited greater loss of skeletal muscle mass and, consequently, a greater reduction in basal metabolic rate. This result necessitates the development of strategies and different nutritional protocols for the elderly, which can minimize this loss of muscle mass during a longer period of hospitalization.

The relationship between LCD and weight loss and its effects on glucose, lipid, and inflammatory parameters are well documented in the scientific literature (8). In this study, the reduction in blood glucose, insulin, glycated hemoglobin, triglycerides, LDL- and total cholesterol, and CRP was significant in both periods of hospitalization. A small reduction in HDL cholesterol was observed only in the short term and was not maintained after 6 months of hospitalization. These results suggest that inpatient treatment improves the metabolic and inflammatory profile of patients with severe obesity, with implications for reducing cardiovascular risk.

Reducing body weight by 5 to 10% improves cardiovascular health, as lipid parameters tend to reduce significantly within this range of weight loss when they are at abnormal levels [23]. The non-significant long-term reduction in HDL cholesterol can be partly explained by the progressive increase in physical activity during hospitalization, associated with improved eating patterns. Therefore, multidisciplinary therapy aimed at lasting changes in lifestyle habits is crucial.

Hepatic transaminase GGT levels reduced significantly throughout both periods of hospitalization. GPT levels increased slightly in the first 3 months but decreased at 6 months. Despite not changing significantly in the first 3 months of hospitalization, the GOT level increased slightly at 6 months. This increase may be because of weight loss during hospitalization; it was within the normal range and without clinical implications.

Fasting blood glucose, insulin, and glycated hemoglobin levels were significantly reduced in this study, indicating an improvement in glycemic control and insulin resistance, corroborating previous reports [24–25]. This improvement in glucose parameters demonstrates that this treatment reduces weight and body fat, achieves positive metabolic profile results, and removes the patient from pre-diabetes classification.

In both periods of hospitalization, inflammatory marker levels, CRP, and ferritin were also reduced significantly. Reducing weight and body fat mass reduces free fatty acid levels that activate the pro-inflammatory cascade. This increases adiponectin levels, reducing CRP production by reducing the release of interleucin-6 by adipose tissue [24–26]. Elevated ferritin is an important marker of chronic inflammation, cardiovascular risk, and insulin resistance and is associated with inflammation in adipose tissue [27–28]. Therefore, the reduction in ferritin and CRP levels observed in this study reflects an improvement in the inflammatory parameters of patients with severe obesity.

A logistic multivariate regression analysis was conducted to understand the factors, measured at admission, associated with the success to reach the median value of fat mass loss percentage at 3 and 6 months of treatment. The model explained approximately 33.5%% and 28.7% of the success to reach such levels of fat mass loss percentage variation after 3 and 6 months of treatment, respectively. Male sex was a significant predictor of the percentage of fat mass loss at both treatment times. Previous reports have indicated greater weight and fat loss in men who underwent interventions to treat obesity [15,29]. A higher percentage of lean mass in men, a greater amount of estrogen produced by women, and differences in insulin resistance between sexes may justify these different responses to treatment [29–30]. Another significant predictor was age; elderly people presented lower chance of reach median values of fat mass loss percentage in both periods of hospitalization, corroborating previous data [21]. Metabolic changes inherent to advancing age, associated with reduced muscle mass and strength [31–32], may explain this negative association with age.

Alcoholism reported on admission was related to a higher chance to reach median percentage of fat mass loss in the first 3 months of hospitalization but not at 6 months. Upon admission, patients stop drinking alcoholic beverages, contributing to a reduction in caloric intake and, consequently, a reduction in fat mass in the short term. Hepatic steatosis was a predictor associated with a higher chance of reach median percentage of fat mass loss at six months of hospitalization. As the liver is one of the main metabolic regulatory organs, hepatic steatosis indicates an altered and compromised metabolism, which seems to respond better to a six months inpatient treatment.

Patients with high CPK levels on admission had a higher percentage of fat mass loss in both periods of hospitalization. CPK is an enzyme in muscle cells, with only small amounts released into the bloodstream [33–36]. However, high body fat levels can generate changes in the cell membrane, increasing circulating CPK levels [37]. As the main source of CPK is muscle tissue, the levels of this enzyme can indicate muscle mass, justifying its association with a better response in the loss of body fat. However, more studies are necessary to verify this association. CRP level was a negative predictor of fat mass loss at patient admission in this study. CRP level is an important indicator of subclinical systemic inflammation in individuals with severe obesity [38]. Therefore, the greater difficulty in losing fat mass in patients with higher CRP levels could be explained by a worse inflammatory profile resulting from more dysfunctional adipose tissue. However, more studies are needed to address this question.

The zinc level below normal range was associated with a lower chance to reach median level of fat mass reduction at 3 but not at 6 months of treatment. Zinc plays an important role in controlling appetite, has anti-inflammatory action, and is involved in the production of hormones associated with energy metabolism, insulin resistance, and diabetes [39–40]. Therefore, the zinc association reported herein could be explained by its protective effects against insulin resistance, chronic inflammation, and hyperglycemia, leading to higher fat mass loss. Its effect loss at 6 months may be because this micronutrient is supplemented during hospitalization, correcting possible deficiencies and their effects in the long term.

## Conclusion

Three and 6 months of inpatient treatment substantially reduced anthropometric measurements in severe obesity, with 6 months yielding better results with approximately 20% reduction in weight, 31% in fat mass, and 6% in the WHR. Furthermore, glucose, lipid, and inflammatory profiles significantly improved in both treatment periods. These results were maintained in elderly and non-elderly patients, men and women, resulting in an improvement in cardiovascular risk and escape from pre-diabetes. This study suggests that VLCD in an inpatient facility with immersive lifestyle changes under multidisciplinary supervision is an alternative and effective intervention for managing severe obesity in the real world.

## Data Availability

All relevant data are within the manuscript and its Supporting Information files.

## Acknowledgment

We thank all the professionals of Hospital da Obesidade for their kind support.

